# Depression, loneliness, and malnutrition among older people in Lalitpur, Nepal: findings from a 2014 community-based cross-sectional study

**DOI:** 10.64898/2026.04.29.26352044

**Authors:** Eebaraj Simkhada, Aerusha Simkhada

## Abstract

**Background:** As Nepal’s older population grows and chronic health conditions become more common, the vulnerability of older adults has emerged as a major public health concern. This study assessed the key aspects of geriatric health—depression, loneliness, and malnutrition, along with the factors associated with depression—among older people living in the Lalitpur district, Nepal.

**Methods:** This study utilized a multistage cluster sampling technique, with probability proportionate to size, to select 360 participants aged 60 and older from the communities in Lalitpur, Nepal. Depression, loneliness, and nutritional status were assessed via the Geriatric Depression Scale (GDS), the Modified Jong-Gierveld scale, and the Mini Nutritional Assessment (MNA) tool, respectively.

Statistical analysis was conducted using complex sample statistics, with the findings presented as percentages. To determine the relationships between variables, chi-square and F-tests were utilized. Furthermore, simple linear regression analysis with wild cluster bootstrapping was employed to identify factors associated with depression, with the results expressed as coefficients accompanied by 95% confidence intervals.

**Results:** In the study population, 27% of the participants had depression, and 6.1% had severe depression. Approximately 67% were experiencing loneliness, and nearly 25% reported severe loneliness. Furthermore, about 48.9% were considered socially isolated. The prevalence of malnutrition and at risk of malnutrition was 13% and 45%, respectively. In the regression analysis adjusted for sex, family structure and marital status, a significant positive association was observed between depression and being female gender (β = 0.11; 95%CI = 0.01 − 0.21), having financial dependency(β = 0.14; 95%CI = 0.00 − 0.27), at risk of malnutrition (β = 0.20; 95%CI = 0.08 − 0.31) being malnourished (β = 0.58; 95%CI = 0.45 − 0.72), having insomnia (β = 0.22; 95%CI = 0.10 − 0.35), having smoking history (β = 0.14; 95%CI = 0.00 − 0.28) and experiencing loneliness (β = 0.34; 95%CI = 0.24 − 0.44).

**Conclusion:** This study offers unique insights into the prevalence of depression, loneliness, and malnutrition among older adults living in Lalitpur district of Nepal, and key factors significantly associated with depressive: female gender, financial dependence on others, experiencing loneliness, having insomnia, smoking history and malnutrition. These findings underscore a critical requirement for comprehensive health screenings and targeted public health strategies to mitigate the primary wellness challenges faced by the aging population in the community.

## Background

Global longevity is on the rise, leading to a significant expansion of the population aged 60 and above [1]. Mirroring this global trend, Nepal is also seeing a steady increase in its older population. Data from the 2011 national census reveald that individuals aged 65 and older accounted for 8.1% of the country’s total population [2].

While society benefits immensely from the contributions of older adults, these vital roles depend heavily on their physical and mental well-being. Conversely, a decline in the health status of this population can lead to broader social and economic challenges [1]

Advancing age is frequently accompanied by a complex interplay of multiple chronic conditions and various physical or mental health hurdles. The rising level of comorbidity intensifies the demand for specialized medical and social support, making difficulty to address the unique requirements of the elderly [3]. Consequently, an older population places significant pressure on healthcare infrastructure [4].

Late-life depression represents a prevalent health concern within the older population. Depression is characterized by persistent sadness, insomnia, guilty feelings, decreased energy, loss of concentration, loss of appetite, less interest in pleasurable activities, increased or decreased psychomotor activity, and suicidal ideation. The diagnosis of major depression is made on the basis of the presence of 5 out of the 9 symptoms. Symptoms must be present over a period of 2 weeks in a person [5].

Significant risk factors for late-life depression include exposure to traumatic events, childhood maltreatment, and the accumulation of stressful life transitions [6]. This vulnerability is further exacerbated by a lack of adequate social and familial support systems, which has been shown to increase depressive symptoms among the older individuals [7].

Research indicates that depression in this population leads to a cascade of adverse outcomes, including a diminished quality of life and functional limitations in daily living. Furthermore, it is associated with deteriorating physical health, cognitive decline, and an increased risk of premature mortality [8]

Similarly, loneliness is a prevalent condition that disproportionately affects the older people. It is a painful, negative, subjective feeling or social pain resulting from a discrepancy between preferred and actual social connections [9].

Human beings are inherently social, requiring ample interpersonal connections to maintain both psychological and physical well-being. However, as individuals age, opportunities for social engagement often contract, heightening their susceptibility to loneliness. This vulnerability is frequently exacerbated by late-life stressors—such as retirement, the death of a spouse, the migration of children, or the onset of disability—all of which can compromise overall health [10]. Furthermore, loneliness and social isolation serve as significant risk factors for a range of severe conditions, including cardiovascular disease, stroke, diabetes, cognitive impairment, dementia, and mental health crises such as depression, anxiety, and suicide [11].

Good social connections are a fundamental pillar of human well-being. While social isolation and loneliness are increasingly recognized as critical issues in geriatric health, these specific social determinants often remain overlooked in broader health policy and research [12].

Additionally, malnutrition is a prevalent condition among the older people, with a disproportionately high incidence in low-income populations. Social determinants—including economic hardship, social isolation, and limited educational attainment—can severely restrict access to adequate nutrition, consequently compromising overall state of health [13].

Undernutrition is a state resulting from a lack of intake or uptake of nutrients that leads to altered body composition and body cell mass. Malnutrition in the elderly is defined as a state of deficient or imbalanced nutrition resulting from insufficient food intake, diminished appetite, sarcopenia (muscle wasting), and involuntary weight loss [3]. This condition is rarely isolated; rather, the nutritional status of an older adult is shaped by a multifaceted interplay of dietary habits, socioeconomic conditions, and physical and psychological health [14].

Malnutrition remains highly prevalent in low-income nations, where approximately 13.5% of the general population was identified as chronically undernourished in 2014. During that same period, data form Nepal indicated that 8.1% of its citizens—approximately 3.6 million people—suffered from undernourishment [15]. However, the prevalence among the older population may be even higher. A study in Nepal utilizing the Mini Nutritional Assessment (MNA) tool found that 31% of older adults were malnourished [17]. This finding underscores the critical severity of nutritional deficiency among the aging population in the Nepalese population.

### Rationale of the study

National health frameworks and initiatives in Nepal have placed less priority to geriatric wellness. Consequently, there remains a significant scarcity of comprehensive data regarding the nutritional status, physical, mental health, and social well-being of the older people. Enhancing our understanding of these determinants of health in old age is crucial for designing effective preventive and therapeutic interventions, ultimately improving the quality of life for seniors within their communities. Lalitpur was one of the area which was experiencing rapid urbanization and demographic transition leading to an increasing proportion of older people in the population. In addition, the urban setting of Lalitpur provided a diverse older population with varying socioeconomic backgrounds, living arrangements, and access to health services. It was an urban poverty area, having a significant proportion of poverty in urban area [18]. Taking these factors in consideration, this location was selected for the study. The critical knowledge gap concerning loneliness, depression, and malnutrition in Nepal has been formally recognized in the national status report on the elderly [4]. In this context, this study aimed to investigate the states of the primary dimensions of geriatric health—specifically depression, loneliness, social connectivity, and malnutrition—among community-dwelling older adults in Lalitpur, Nepal.

## Methods

### Study designs and settings

This was a cross-sectional, population-based study among older people aged 60 years or older in 13 VDC (Village Development Committee) of Lalitpur, Nepal. For this study, we used multistage cluster sampling with a probability proportional to size method. This method is widely used in health research, especially in national health surveys such as demographic and health surveys. It has an established methodology, accepted statistical validity, and easy comparability with other studies. Probability proportional to size ensures that a person living in a high-density area has the same individual probability of being included in the survey as someone in a low population density area. It mathematically balances the size of the cluster against the likelihood of selection.

For the sampling purpose, this research utilized the administrative framework predating Nepal’s 2017 federal restructuring. Specifically, geographical clusters were identified based on the traditional division of Districts, Village Development Committees (VDCs), and Municipalities. At the time of data collection, the transition to the new federal system had not yet been implemented. Following the national reorganization, the VDCs included in the study areas were officially integrated into larger municipalities [19].

Data collection for the study was conducted from September 20, 2014 to November 7, 2014.

Despite the time elapsed between data collection and the publication of this report, this study offers vital, foundational insights into depression, loneliness, social isolation, and malnutrition among the older adults in Nepal’s Lalitpur district. These dimensions of geriatric health were unexamined previously within this particular community. These findings serve as a critical reference for policymakers and program planners designing geriatric interventions. Furthermore, this data establishes a baseline status, enabling future researchers to track and evaluate longitudinal changes in the wellbeing of the aging population.

### Study population and sampling

The study population included elderly people residing in the surrounding VDCs of Lalitpur metropolitan city in Lalitpur district, Nepal. The district is divided into 3 electoral constituencies. Densely populated electoral constituencies no. 2 and 3, consisting of 13 VDCs, were selected purposively. Five VDCs (Lele, Siddhipur, Thaiba, Saibu, and Khokana) were selected randomly from the 13 VDCs. Each VDC is divided into 9 wards, and each ward is taken as a cluster. In total, from 45 clusters, 30 clusters were selected according to probability proportionate to size. A sample of 12 elderly individuals was selected randomly from each cluster. Older person from each household was selected on the basis of the willingness to participate and the availability of the respondent.

The inclusion criteria of the study were participants 60 years or older, not being blind and/or deaf, consenting to participate in the study, not being bed-bound due to poor health, and not having severe mental illness.

The sample size was determined according to the prevalence of malnutrition among home-living older people, and the prevalence of depression was also considered.

The prevalence of depression in elderly individuals was reported to be 29.7% in a study [20] in Kathmandu, Nepal. Another study conducted in Pokhara, Nepal, reported a 26.7% prevalence of depression [21].

A review of the literature on the MNA (Mini Nutritional Assessment) [22] for the prevalence of malnutrition among the community-dwelling elderly population, revealed 2% malnutrition and 24% at risk of malnutrition in developed countries. A study from Bangladesh [23] reported 26% malnutrition among rural elderly people in Bangladesh using the MNA tool to assess nutritional status. A study carried out near Kathmandu Valley using the MNA revealed that 31% of elderly people were malnourished and that 51% were at risk of malnutrition [17]. The prevalence of 31% was taken on the basis of the results of different studies.

#### Sample Size Calculation

Overall, 360 older people were included in the study. The total sample size was calculated by considering prevalence of malnutrition among older adults as 31% with 7% margin of error, 2% design effect and 8% non response rate. The detail calculation is presented below.

n= Z²x p x q / e² x DEFF

= (1.96)²x 0.31x 0.69/ (0.07)² x DEFF

= 167x 2

= 334

Non response rate of 8% is added,

= 334 +26

= 360 (final sample size) Where,

n= sample size

Z= 1.96 at 95% Confidence level

p= prevalence of older population malnutrition (31%)

q= 1-p

e= margin of error, 7%

DEFF= design effect, 2.0

A multicomponent structured questionnaire including assessment tools was used to collect the data in this study. The questionnaire was translated from English into Nepali and then back-translated into English by two people fluent in both languages. Pretesting of the questionnaire was performed on a sample of 30 elderly people to identify the feasibility of the questionnaire. In accordance with the feedback, the questionnaire was revised with minor grammatical changes for the clarity.

### Validity and reliability

Validated instruments were used. Probing questions were asked. The MNA tool was not validated at the time of the study, however later it was validated in a study among older people in Nepal and had shown good reliability [24], and the GDS-15 scale has been validated in the Nepali language [25]. The modified Jong-Gierveld Loneliness Scale has been used in many countries, including in national surveys [26].

Reliability was assured through the translation and retranslation of tools (English-Nepali-English). The study tools and instruments were pretested at the Tikathali VDC. The training of the enumerators was conducted.

### Study variables

#### Dependent variables

Depression, and loneliness were considered variables of interest.

#### Depression

The short version of Yesavage’s Geriatric Depression Scale [27], which includes 15 items, was used. It is a widely used instrument to assess the affective state of elderly people. Each item is dichotomized as “yes” or “no” and is scored according to the individual’s feeling over the past week. The score ranges from 0-15. The cutoff points were 0-5 for normal, 6-10 for mild depression, and ≥ 11 for severe depression.

#### Loneliness and social isolation

The modified Jong-Gierveld loneliness scale, as described by Wilson et al. [28] was used to evaluate the loneliness of the study population. This scale is based on 5 questions: “I experience a general sense of emptiness,” “I miss having people around,” “I feel like I don’t have enough friends,” “I often feel abandoned,” and “I miss having a truly good friend.” Answers were “yes,” coded as 1, and “no,” coded as 0. The score ranges from 0-5, and according to the author, the higher the value is, the greater the level of loneliness in an individual.

The Lubben Social Network Scale 6 [29] was used to assess the social isolation of the individuals. This 6-item scale included the following questions: “How many relatives do you see or hear from at least once a month?” “How many relatives do you feel at ease with that you can talk about private matters?” and “How many relatives do you feel close to such that you could call on them for help?” These same questions were repeated by replacing the word “relatives” with the word “friends.” The answers were recorded as follows: none = 0, one = 1, two = 2, three or four = 3, five through eight = 4, and nine or more = 5. The score ranged from 0-30, and a score < 12 was considered a risk of social isolation.

#### Independent Variables

Variables such as sex, age, education, marital status, family structure, main occupation, ethnicity, financial dependence on children, living arrangements (living alone or living with others), daily drug intake, comorbidities, recent hospitalization, insomnia, chronic pain, current smoking, nutritional status, and physical exercise were considered independent variables.

#### Nutritional status

The MNA is used as a tool for the assessment of nutritional status in older people. This assessment tool is widely used in aged people. People having malnutrition and at risk of malnutrition in MNA score were considered as having poor nutritional status.

The MNA includes 18 questions grouped into 4 parts, which are assessed simultaneously.

1. Weight and height, weight loss, and arm and calf circumferences were measured.
2. General assessments such as lifestyle, medication, mobility, and the presence of signs of depression or dementia were recorded.
3. Dietary assessment was performed, recording the number of meals, food and fluid intake, and autonomy of feeding.
4. Subjective assessments such as self-perception of health and nutrition were documented.

Each answer had a score, and the maximum MNA score was 30. It is a nutritional status assessment tool with a reliable scale and clearly defined thresholds.

The MNA was first translated into Nepali carefully to preserve the original meaning of the MNA questions and then revised by the researcher and field interviewers. The MNA score of each participant was calculated, and scores were categorized as < 17 points for malnutrition, 17-23.5 points for nutritional risk, and ≥ 24 points for being well nourished. A score < 24 was considered poor nutritional status.

An electronic digital scale from Microlife, Switzerland (model no. WS 50), was used to measure weight. Body weight was measured in light clothes and no shoes to the nearest 0.1 kg, and height was measured via a measuring tape mounted on a wall to the nearest 0.5 cm. The measurements were taken while the participants were standing without shoes, with their feet placed together with the heels, buttocks, shoulders, and backs of the head touching the wall and with the head facing straight forward. The mid upper arm circumference was measured with a nonelastic tape on the relaxed arm. Calf circumference was measured while the individual was sitting with a flexed knee at a 90-degree angle with a nonelastic tape on the thickest part of the undressed calf.

Socioeconomic status was assessed by a weighted wealth index [30]. Household wealth was constructed via household asset data and household characteristics. The index assessed housing characteristics, ownership of agricultural land, and household assets in the house. The availability and use of drinking water for the household, the use of fuel for cooking, and the use of a toilet were also assessed by the index. The total wealth index score was then divided into five categories: lowest, second, middle, fourth, and highest income groups. Household wealth quintiles were considered very poor (lowest wealth quintile), poor (second wealth quintile), middle (third wealth quintile), rich (fourth wealth quintile), and very rich (fifth wealth quintile) in the study.

All the independent variables of the study are presented in Table 1.

**Table 1.** Independent variables of the study and definitions.

| Variable | Definition and categories |
| --- | --- |
| Age | Age in completed years, categorized as 60-69, 70-79, 80+ |
| Sex | Male, Female |
| Education | Respondent's highest level of education; categorized as:<br>Illiterate: not able to read and write<br>Literate: Having informal education and able to read and write<br>Primary education: formal education, one class to five classes<br>Secondary and above: Formal education, six classes and above |
| Ethnicity | Brahmin/Chettri, Newar, other Janajati (Magar, Tamang, Gurung), and Dalit |
| Family structure | Type of the family structure was categorized into 3 groups:<br>Nuclear, joint and extended family. |
| Marital status | Married, Never Married, Divorced, Widowed |
| Occupation | Main occupation was recorded based on current occupation as farmer, laborer, self-employed/job, and housework/no work. |
| Household Wealth quintels | Poorest, Poor, Middle, Rich, Very Rich |
| Financial dependency | “totally dependent,” “partially dependent,” and “independent” |
| Living condition | Classified as living alone or living with family. |
| Smoking | Assessed by asking “yes” and “no” questions regarding whether the elderly person was currently smoking or not. Current smoking was defined as the participant having smoked within the past month. |
| Daily drug intake | Taking prescribed drug daily documented as one, two, three drugs, and three or more. |
| Exercise | Assessed by asking if the individual had been exercising daily for |
|  | 30 minutes for over a year and coded as Yes or No. |
| Comorbidity | Self-reported comorbidities were recorded based on physician-diagnosed chronic diseases. Responses were again recorded in categories: no chronic illness, one to two chronic illnesses, and three or more chronic illnesses. |
| Chronic pain | Chronic pain was defined as participants feeling pain for at least 3 months, as determined by asking a yes/no question. |
| Insomnia | Responses were categorized into two: no or occasionally, and often or always. |
| Recent Hospitalization | Recent hospitalization, defined as hospitalization within 1 year, was recorded as yes or no. |

### Statistical analyses

#### Data Management and Analysis

Data editing and coding were performed, and data entry was performed in EpiData 3.1. Data analyses were carried out via the Statistical Package for the Social Sciences (SPSS) software, version 21.0. In order to obtain representative results from the study, sample weights were calculated based on the sample design and complex sample analysis was performed. Percentages were used to present nominal variables, whereas chi-square and F tests were performed to assess bivariate associations between variables. To evaluate the observed associations between depression and other variable, a linear regression model, was used in Stata (17.0, StataCorp LLC, College Station, TX). Taking depression as the dependent variable, adjustments were made for sex, family structure, and marital status. Additionally, the standard errors were clustered to account for the 30 specific clusters within the data. To address non-normally distributed data and the relatively small sample size, wild cluster bootstrap procedure of David Roodman [31] with 2,999 replications were applied to generate p-values, robust estimates of correlates and their 95% confidence intervals. The statistical significance was set at a p-value < 0.05.

### Ethical consideration

Ethical approval was obtained from the institutional review board of the Institute of Medicine (IOM), Maharajgunj (Ref no. 75(6-11-E)071/072). The enumerators were selected and trained for documentation, record keeping, and data collection. Prior to the interview, written consent was obtained from all participants, and thumb impressions were obtained from illiterate participants. The participants were fully informed of their rights to decline or withdraw from participation in the study if desired. In the case of an inability to communicate effectively due to language or speech leading to difficulty in understanding, the help of a family member was obtained. Data privacy was maintained in the study.

## Results

### Sociodemographic characteristics of the study population

The research involved 360 older participants, comprising 183 females (50.8%) and 177 males (49.2%). Participants ranged in age from 60 to 84, maintaining an average age of 71.2 years. Regarding ethnic composition, the group was predominantly Newar (68%), while Dalits represented 1.7% of the cohort.

The demographic profile reveals that 67.8% of the participants had no formal education. Living arrangements were largely communal, with 85.8% of the elderly residing in joint family households. In terms of occupation, the majority (71.7%) were engaged in farming, while the remaining participants were evenly split between those in business or employment (14.2%) and those who were unemployed or focused on domestic duties (14.2%).

The bivariate relationships between the independent variables and the dependent variables were examined. Table 2 presents the detailed characteristics of the study participants according to those who were depressed and not depressed, and those with loneliness and without loneliness.

**Table 2.** Sociodemographic characteristics according to depression, loneliness and nutritional status.

| Variables | Total | No | Depression | P | No | Loneliness | P |
| --- | --- | --- | --- | --- | --- | --- | --- |

|  | **frequency**<br>**n=360** | **depression**<br>**n=263** | **n=97** |  | **loneliness**<br>**n=119** | **n=241** |  |
| --- | --- | --- | --- | --- | --- | --- | --- |
|  | n(%) | n (%) | n (%) |  | n (%) | n (%) |  |
| **Age class** |  |  |  | 0.887 |  |  | 0.180 |
| 60-70 | 200(55.6) | 144(54.8) | 56(57.7) |  | 73(61.3) | 127(52.7) |  |
| 71-80 | 101(28.1) | 76(28.9) | 25(25.8) |  | 33(27.7) | 68(28.2) |  |
| >80 | 59(16.4) | 43 (16.3) | 16 (16.5) |  | 132(10.9) | 46(19.1) |  |
| **Gender** |  |  |  | 0.022 |  |  | 0.271 |
| Male | 177(49.2) | 140 (53.2) | 123(46.8) |  | 64(53.8) | 113(46.9) |  |
| Female | 183(50.8) | 37(38.1) | 60(61.9) |  | 55(46.2) | 128(53.1) |  |
| **Ethnicity** |  |  |  | 0.134 |  |  | 0.742 |
| Brhamin/Chettri | 97(26.9) | 73(27.8) | 24(24.7) |  | 31(26.1) | 66(27.4) |  |
| Newar | 243(67.5) | 179(68.1) | 64(66.0) |  | 83(69.7) | 160(66.4) |  |
| Janjati (other) | 14(3.9) | 11(4.2) | 3(3.1) |  | 5(4.2) | 9(3.7) |  |
| Dalit | 6(1.7) | 0(0.0) | 6(6.2) |  | 0(0.0) | 6(2.5) |  |
| **Living status** |  |  |  | 0.848 |  |  | 0.260 |
| Living alone | 3(0.8) | 2(0.8) | 1(1.0) |  | 0(0.0) | 3(1.2) |  |
| Living together | 357(99.2) | 261(99.2) | 96(99.0) |  | 119(100.0) | 238(98.8) |  |
| **Marital status** |  |  |  | 0.320 |  |  | 0.536 |
| Married | 212(58.9) | 159(60.5) | 53(54.6) |  | 73(61.3) | 139(57.7) |  |
| Widowed | 148(41.1) | 104(39.5) | 44(45.4) |  | 46(38.7) | 102(42.3) |  |
| **Education** |  |  |  | 0.507 |  |  | 0.837 |
| No education | 244(67.8) | 171(65.0) | 73(75.3) |  | 79(66.4) | 165(68.5) |  |
| Literate | 52(14.4) | 42(16.0) | 10 (10.3) |  | 20(16.8) | 32(13.3) |  |
| Primary | 25(6.9) | 16(6.1) | 9(9.3) |  | 9(7.6) | 16(6.6) |  |
| Secondary and above | 39(10.8) | 34(12.9) | 5(5.2) |  | 11(9.2) | 28(11.6) |  |
| **Wealth quintile** |  |  |  | 0.003 |  |  | 0.657 |
| Lowest | 72(20) | 41(15.6) | 31(32.0) |  | 21(17.6) | 51(21.2) |  |
| Second | 72(20) | 52(19.8) | 20(20.6) |  | 30(25.2) | 42(17.4) |  |
| Middle | 72(20) | 57(21.7) | 15(15.5) |  | 22(18.5) | 50(20.7) |  |
| Fourth | 72(20) | 61(23.2) | 11(11.3) |  | 24(20.2) | 48(19.9) |  |
| Highest | 72(20) | 52(19.8) | 20(20.6) |  | 22(18.5) | 50(20.7) |  |
| **Financial dependency** |  |  |  | 0.028 |  |  | 0.747 |
| No | 92(25.6) | 78(29.7) | 14(14.4) |  | 29(24.4) | 63(26.1) |  |
| partially/Total | 268(74.4) | 185(70.3) | 83(85.6) |  | 90(75.6) | 178(73.9) |  |
| **Main occupation** |  |  |  | 0.501 |  |  | 0.422 |
| Agriculture | 258(71.7) | 193(73.4) | 65(67.0) |  | 91(76.5) | 167(69.3) |  |
| Employed | 51(14.2) | 37(14.1) | 14(14.4) |  | 16(13.4) | 35(14.5) |  |
| Housework | 51(14.2) | 33(12.5) | 18(18.6) |  | 12(10.1) | 39(16.2) |  |
| p values are from adjusted F, which is a variant of the second-order Rao-Scott adjusted Chi square test |  |  |  |  |  |  |  |

Table 3 presents the results of the comparison of the health characteristics between those who were depressed and not depressed, those with loneliness and without loneliness, and those with normal nutrition and poor nutrition in the study population.

**Table 3.** Sample health characteristics according to depression, nutrition and loneliness.

| **Variables** | **No**<br>**depression**<br>**n=263** | **Depression**<br>**n=97** | **P** | **No**<br>**loneliness**<br>**n=119** | **Loneliness**<br>**n=241** | **P** | **Normal**<br>**nutrition**<br>**n=151** | **Poor**<br>**nutrition**<br>**n=209** | **P** |
| --- | --- | --- | --- | --- | --- | --- | --- | --- | --- |
|  | n (%) | n (%) |  | n (%) | n (%) |  | n (%) | n (%) |  |
| **Age class** |  |  | 0.887 |  |  | 0.180 |  |  | 0.057 |
| 60-70 | 144(54.8) | 56(57.7) |  | 73(61.3) | 127(52.7) |  | 93(61.6) | 107(51.2) |  |
| 71-80 | 76(28.9) | 25(25.8) |  | 33(27.7) | 68(28.2) |  | 42(27.8) | 59(28.2) |  |
| >80 | 43 (16.3) | 16 (16.5) |  | 132(10.9) | 46(19.1) |  | 16(10.6) | 43(20.6) |  |
| **Gender** |  |  | 0.022 |  |  | 0.271 |  |  | 0.350 |
| Male | 140 (53.2) | 123(46.8) |  | 64(53.8) | 113(46.9) |  | 79(46.9) | 98(46.9) |  |
| Female | 37(38.1) | 60(61.9) |  | 55(46.2) | 128(53.1) |  | 72(47.7) | 111(53.1) |  |
| **Ethnicity** |  |  | 0.134 |  |  | 0.742 |  |  | 0.113 |
| Brhamin/Chetri | 73(27.8) | 24(24.7) |  | 31(26.1) | 66(27.4) |  | 35(23.2) | 62(29.7) |  |
| Newar | 179(68.1) | 64(66.0) |  | 83(69.7) | 160(66.4) |  | 111(73.5) | 132(63.2) |  |
| Janjati (other) | 11(4.2) | 3(3.1) |  | 5(4.2) | 9(3.7) |  | 3(2.0) | 11(5.3) |  |
| Dalit | 0(0.0) | 6(6.2) |  | 0(0.0) | 6(2.5) |  | 2(1.3) | 4(1.9) |  |
| **Living status** |  |  | 0.848 |  |  | 0.260 |  |  | 0.715 |
| Living alone | 2(0.8) | 1(1.0) |  | 0(0.0) | 3(1.2) |  | 1(0.7) | 2(1.0) |  |
| Living together | 261(99.2) | 96(99.0) |  | 119(100.0) | 238(98.8) |  | 150(99.3) | 207(99.0) |  |
| **Marital status** |  |  | 0.320 |  |  | 0.536 |  |  | 0.086 |
| Married | 159(60.5) | 53(54.6) |  | 73(61.3) | 139(57.7) |  | 96(63.6) | 116(55.5) |  |
| Widowed | 104(39.5) | 44(45.4) |  | 46(38.7) | 102(42.3) |  | 55(36.4) | 93(44.5) |  |
| **Education** |  |  | 0.507 |  |  | 0.837 |  |  | 0.152 |
| No education | 171(65.0) | 73(75.3) |  | 79(66.4) | 165(68.5) |  | 98(64.9) | 146(69.9) |  |
| Literate | 42(16.0) | 10 (10.3) |  | 20(16.8) | 32(13.3) |  | 18(11.9) | 34(16.3) |  |
| Primary | 16(6.1) | 9(9.3) |  | 9(7.6) | 16(6.6) |  | 11(7.3) | 14(6.7) |  |
| Secondary and above | 34(12.9) | 5(5.2) |  | 11(9.2) | 28(11.6) |  | 24(15.9) | 15(7.2) |  |
| **Wealth quintile** |  |  | 0.003 |  |  | 0.657 |  |  | 0.085 |
| Lowest |  |  |  |  |  |  |  |  |  |
| Second | 41(15.6) | 31(32.0) |  | 21(17.6) | 51(21.2) |  | 31(20.5) | 41(19.6) |  |
| Middle | 52(19.8) | 20(20.6) |  | 30(25.2) | 42(17.4) |  | 30(19.9) | 42(20.1) |  |
| Fourth | 57(21.7) | 15(15.5) |  | 22(18.5) | 50(20.7) |  | 34(22.5) | 38(18.2) |  |
| Highest | 61(23.2) | 11(11.3) |  | 24(20.2) | 48(19.9) |  | 35(23.2) | 37(17.7) |  |
|  | 52(19.8) | 20(20.6) |  | 22(18.5) | 50(20.7) |  | 21(13.9) | 51(24.4) |  |
| **Financial dependency** |  |  | 0.028 |  |  | 0.747 |  |  | 0.029 |
| No | 78(29.7) | 14(14.4) |  | 29(24.4) | 63(26.1) |  | 49(32.5) | 43(20.6) |  |
| partially/Total | 185(70.3) | 83(85.6) |  | 90(75.6) | 178(73.9) |  | 102(67.5) | 166(79.4) |  |
| **Main occupation** |  |  | 0.501 |  |  | 0.422 |  |  | 0.920 |
| Agriculture | 193(73.4) | 65(67.0) |  | 91(76.5) | 167(69.3) |  | 107(70.9) | 151(72.2) |  |
| Employed | 37(14.1) | 14(14.4) |  | 16(13.4) | 35(14.5) |  | 22(14.6) | 29(13.9) |  |
| Housework | 33(12.5) | 18(18.6) |  | 12(10.1) | 39(16.2) |  | 22(14.6) | 29(13.9) |  |
| p values are from adjusted F, which is a variant of the second-order Rao-Scott adjusted Chi square test. |  |  |  |  |  |  |  |  |  |

In terms of health status, nearly 59% of the elderly participants reported living with one or two chronic conditions, while 12.2% used a daily regimen of more than three prescribed medications. Regarding lifestyle habits, approximately one-third (34%) were current tobacco smokers, and one-quarter (25%) engaged in regular physical exercise.

Mental health findings revealed that approximately 27% of the cohort exhibited symptoms of depression. Specifically, 20.8% of the population experienced mild to moderate depression, while 6.1% suffered from severe depressive symptoms.

Based on the modified Jong-Gierveld Loneliness Scale, approximately 67% of the participants reported experiencing loneliness. Furthermore, nearly half of the population (48.9%) was classified as socially isolated, defined by a Luben score of less than 12. Additionally, nutritional assessments using the MNA revealed that 13.1% of the study population was malnourished (MNA score < 17).

Table 4 shows the sample health characteristics between those who were depressed and not depressed, those with loneliness and without loneliness, and those with normal nutrition and poor nutrition in the study population.

**Table 4.** Sample health characteristics according to depression, loneliness and nutritional status.

| **Variables** | **No depression**<br>**n=263** | **Depression**<br>**n=97** | **P** | **No loneliness**<br>**n=119** | **Loneliness**<br>**n=241** | **P** | **Normal nutrition**<br>**n=151** | **Poor nutrition**<br>**n=209** | **P** |
| --- | --- | --- | --- | --- | --- | --- | --- | --- | --- |
|  | n (%) | n (%) |  | n (%) | n (%) |  | n (%) | n (%) |  |
| **Depression** |  |  |  |  |  | <0.001 |  |  | <0.001 |
| Yes |  |  |  | 4(3.4) | 93(38.6) |  | 15(9.9) | 82(39.2) |  |
| No |  |  |  | 115(96.6) | 148(61.4) |  | 136(90.1) | 127(60.8) |  |
| **MNA** |  |  | <0.001 |  |  | <0.001 |  |  |  |
| Normal | 136(51.7) | 15(15.5) |  | 69(58.0) | 82(34.0) |  |  |  |  |
| Risk of-Malnutrition | 112(42.6) | 50(51.5) |  | 45(37.8) | 117(48.5) |  |  |  |  |
| Malnutrition | 15(5.7) | 32(33.0) |  | 5(4.2) | 42(17.4) |  |  |  |  |
| **Social isolation score** |  |  | 0.293 |  |  |  |  |  | 0.039 |
| <12 | 123(46.8) | 53(54.6) |  |  |  |  | 64(42.4) | 112(53.6) |  |
| ≥ 12 | 140(53.2) | 44(45.4) |  |  |  |  | 87(57.6) | 97(46.4) |  |
| **Loneliness score** |  |  | <0.001 |  |  |  |  |  | <0.001 |
| 0-1 | 115(43.7) | 4(4.1) |  |  |  |  | 69(45.7) | 50(23.9) |  |
| 2-5 | 148(56.3) | 93(95.9) |  |  |  |  | 82(54.3) | 159(76.1) |  |
| **Number of diseases** |  |  | 0.462 |  |  | 0.444 |  |  | 0.238 |
| No diseases | 246(93.5) | 89(91.8) |  | 108(90.8) | 227(94.2) |  | 145(96.0) | 190(90.9) |  |
| ≤ 2 diseases | 10(3.8) | 3(3.1) |  | 6 (5.0) | 7(2.9) |  | 4(2.6) | 9(4.3) |  |
| > 2 diseases | 7(2.7) | 5(5.2) |  | 5(4.2) | 7(2.9) |  | 2(1.3) | 10(4.8) |  |
| **Daily drug intake** |  |  | 0.617 |  |  | 0.734 |  |  | 0.262 |
| No drug | 128(48.7) | 42(43.3) |  | 52(43.7) | 118(49.0) |  | 78(51.7) | 92(44.0) |  |
| 1-3 drugs | 106 (40.3) | 40(41.2) |  | 52(43.7) | 94(39.0) |  | 60(39.7)) | 86(41.1) |  |
| >3 drugs | 29(11.0) | 15(15.5) |  | 15(12.6) | 29(12.0) |  | 13(8.6) | 31(14.8) |  |
| **Insomnia** |  |  | <0.001 |  |  | 0.003 |  |  | <0.001 |
| Always/Often | 58(22.1) | 45(46.4) |  | 21(17.6) | 82(34.0) |  | 23(15.2) | 80(38.3) |  |
| Occasionally/ | 205(77.9) | 52(53.6) |  | 98(82.4) | 159(66.0) |  | 128(84.8) | 129(61.7) |  |
| No |  |  |  |  |  |  |  |  |  |
| **Chronic pain** |  |  | 0.025 |  |  | 0.859 |  |  | 0.038 |
| No | 225(85.6) | 68(70.1) |  | 96(80.7) | 197(81.7) |  | 131(86.8) | 162(77.5) |  |
| Yes | 38(14.4) | 29(29.9) |  | 23(19.3) | 44(18.3) |  | 20(13.2) | 47(22.5) |  |
| **Smoking** |  |  | 0.040 |  |  | 0.133 |  |  | 0.094 |
| No | 185(70.3) | 53(54.6) |  | 87(73.1) | 151(62.7) |  | 109(72.2) | 129(61.7) |  |
| Yes | 78(29.7) | 44(45.4) |  | 32(26.9) | 90(37.3) |  | 42(27.8) | 80(38.3) |  |
| **Exercise** |  |  | 0.952 |  |  | 0.738 |  |  | 0.002 |
| No | 197(74.9) | 73(75.3) |  | 88(73.9) | 182(75.5) |  | 100(66.2) | 170(81.3) |  |
| Yes | 66(25.1) | 24(24.7) |  | 31(26.1) | 59(24.5) |  | 51(33.8) | 39(18.7) |  |
| **Hospitalized** |  |  | 0.988 |  |  | 0.109 |  |  | 0.013 |
| No | 228(86.7) | 84(86.6) |  | 98(82.4) | 214(88.8) |  | 137(90.7) | 175(83.7) |  |
| Yes | 35(13.3) | 13(13.4) |  | 21(17.6) | 27(11.2) |  | 14(9.3) | 34(16.3) |  |
| p values are from adjusted F, which is a variant of the second-order Rao-Scott adjusted Chi square test. |  |  |  |  |  |  |  |  |  |

In the bivariate analysis of participant characteristics relative to their mental health status revealed that several factors were significantly associated with depression. These included gender and ethnicity, as well as socioeconomic markers like wealth quintile and financial dependency.

Additionally, smoking, insomnia, chronic pain, malnutrition, and loneliness showed a significant association with depression. Significantly higher frequencies of depression were found in the following categories: age 60-70 years (57.6%), female sex (61.9%), Newar caste (66.0%), illiterate (75.3%), financially dependent (85.6%), agricultural occupation (67.0%), at risk of malnutrition (51.5%), malnutrition (33.0%), and loneliness (95.9%).

Furthermore, when loneliness was compared with people not having loneliness, higher frequencies of loneliness were recorded among participants aged 60-70 years, females, the Newar caste, the illiterate group, those who were financially dependent, those with agriculture as their occupation, and those at risk of malnutrition.

### Depression and its correlates

After adjusting for sex, marital status, and family structure, the regression analysis revealed that several factors were significantly associated to depression (Table 5). Specifically, being female, financially dependent, and having a history of smoking showed positive associations. Significant positive correlations were found among individuals who were experiencing loneliness (β = 0.34; 95%CI = 0.24 − 0.44), suffered from insomnia (β = 0.22; 95%CI = 0.10 − 0.35), and were malnourished (β = 0.58; 95%CI = 0.45 − 0.72) or at risk of malnutrition (β = 0.20; 95%CI = 0.08 − 0.31).

**Table 5.**
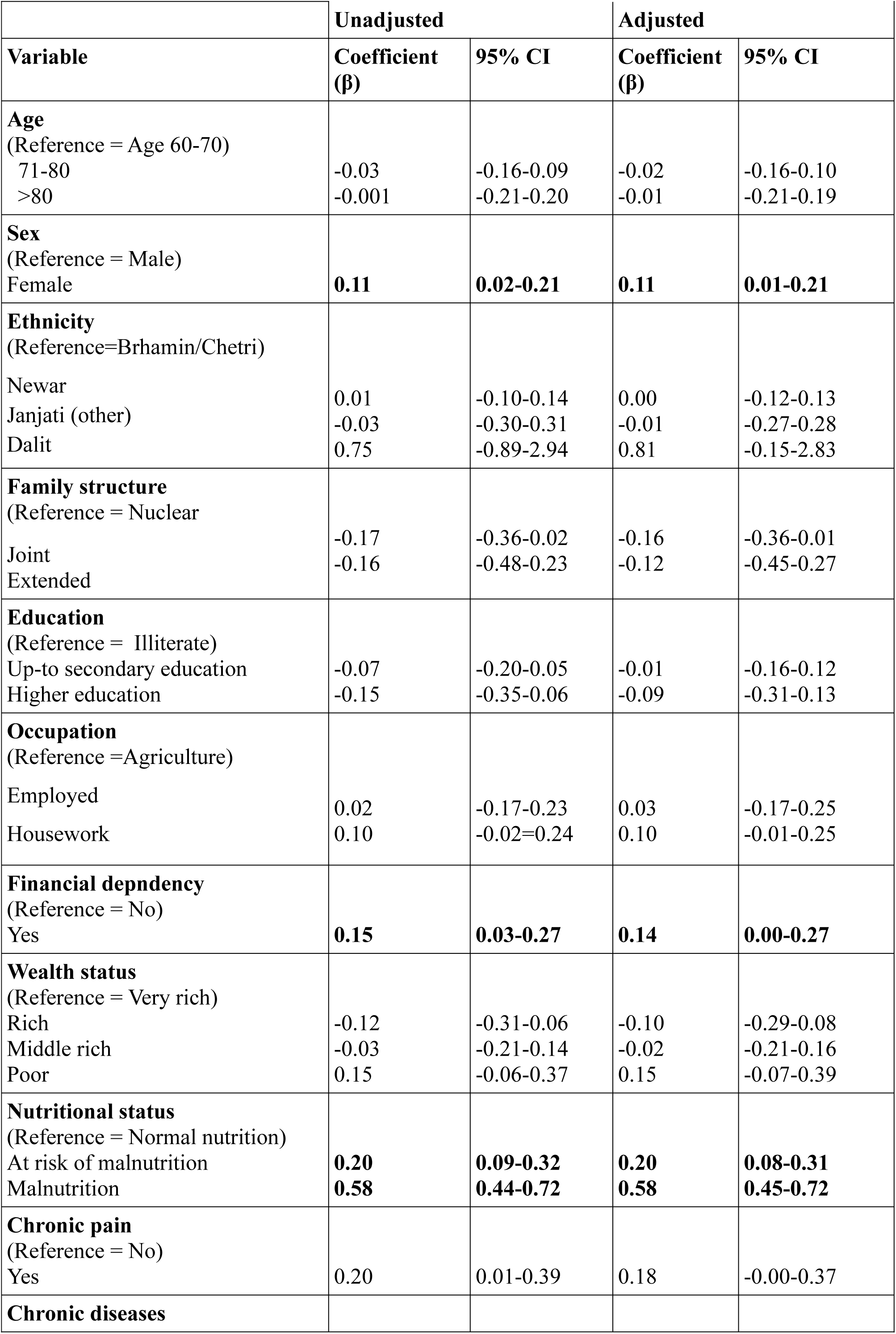
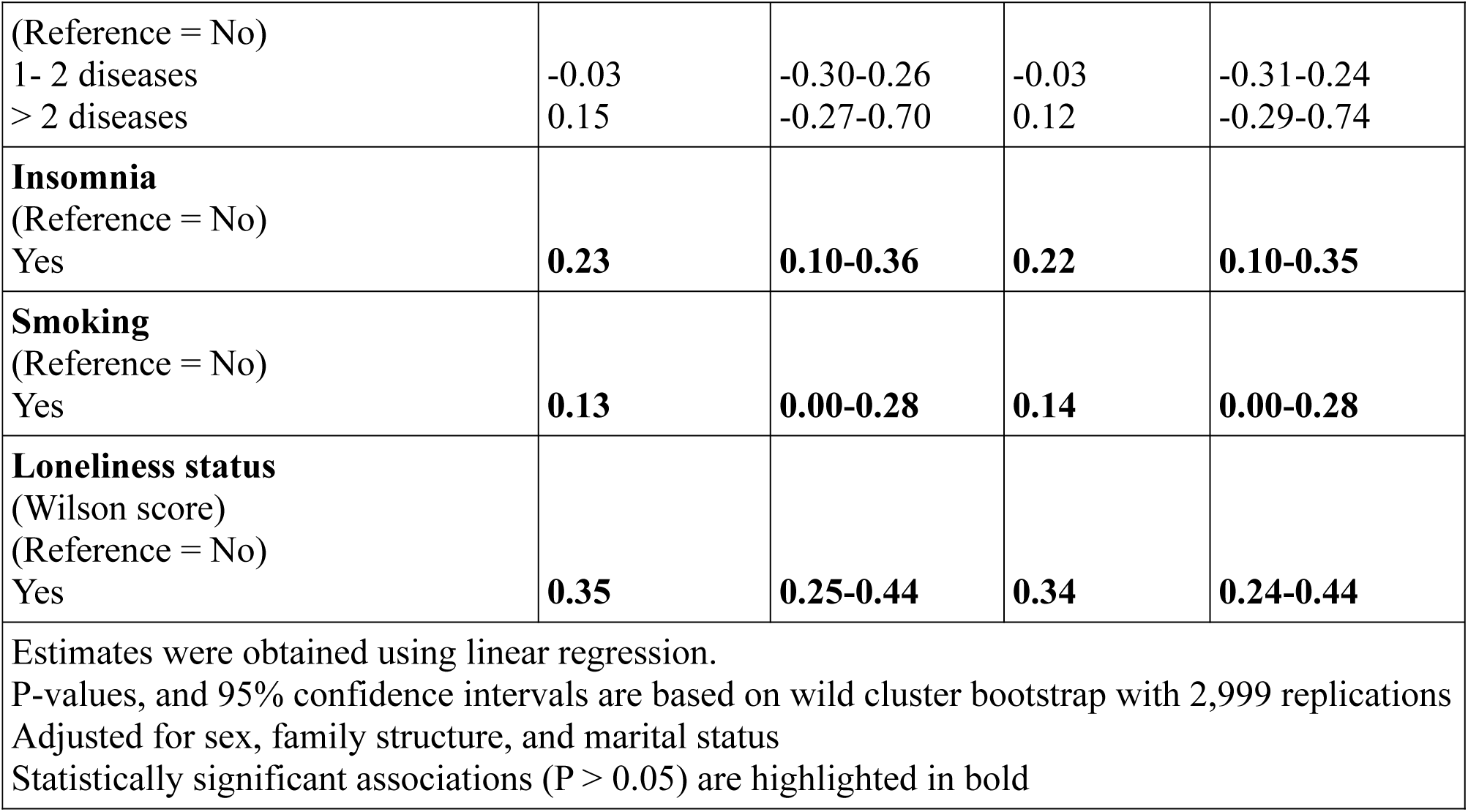
Factors associated with depression among the study population.

## Discussion

This study investigated the prevalence of depression, loneliness, and malnutrition, while also examining the factors associated with depression among older people in Nepal’s Lalitpur district. The findings highlight a concerningly high prevalence of all three conditions within this community. Specifically, being female, financially dependent, and experiencing insomnia and being a smoker were associated with depression. Furthermore, loneliness, malnutrition, and being at risk of malnutrition were all significantly linked to depression in this population. Mental health issues were widespread among the older adults, with more than one fourth of community-dwelling seniors screening positive for depression via the GDS-15 tool.

The study also highlighted a significant burden of subjective loneliness. Specifically, one-fourth of the participants experienced severe loneliness, defined by a score exceeding 2 on the modified Jong-Gierveld scale.

Our study utilized the modified Jong-Gierveld scale to evaluate loneliness, yielding results that align with existing literature. For instance, a study in Lebanon by Beulos et al. [26], which employed the same instrument, identified an 18.5% prevalence of severe loneliness among community-dwelling seniors. In Nepal some of the studies utilized UCLA 3-item Loneliness Scale for the assessment of loneliness in older individuals [9, 16]. The de Jong Gierveld version and UCLA 3-item Loneliness Scale are recognized for their robust psychometric properties, demonstrating high internal consistency, reliable test-retest results, and strong convergent and discriminant validity [32].

While definitive causes remain unproven, factors such as modernization, industrialization, technological advancement, and secularization are frequently linked to rising levels of social isolation and loneliness among the elderly [33]. Beyond these societal shifts, physical health also plays a critical role; the presence of comorbidity and overall poor physical health can heighten the risk of isolation. Similarly, mental health challenges—particularly depression—and sensory deficits, such as vision or hearing loss, significantly contribute to an individual’s sense of loneliness and social withdrawal [10].

In the regression analysis, the current study revealed that experiencing loneliness was associated with depression in the older population. This can be explained by the fact that loneliness is related to physical and cognitive decline in elderly individuals. This could lead to the development of negative beliefs, which are associated with depression. Previous studies [34.35], have demonstrated the link between loneliness, depression, and an increased risk of malnutrition. Loneliness is a modifiable risk factor for depression [36]. However, the reverse causality of depression causing loneliness is also well established. The cognitive discrepancy theory, indicates that depressive symptoms affect cognitive processes and influence how individuals evaluate whether their social interactions are adequate [37].

Social disconnection is prevalent in all regions and all age groups. Its consequences are severe and under-recognized, impacting physical and mental health, well-being, the economy, and wider society. Social isolation and loneliness have serious impacts on health, increasing the risk of cardiovascular disease, stroke, diabetes, dementia, depression, anxiety, and suicide [12].

Its widespread occurrence and severe consequences make it a serious global public health issue [33]. Therefore, public health interventions are critical to prevent loneliness and address the mental health of the aged population.

The present study revealed that more than half of the study population were in poor nutritional state, as measured by the MNA among older individuals. Poor nutritional status, was defined as combined malnutrition and at risk of malnutrition in the study. Hence, our study revealed a lower rate of malnutrition than a previous study [17], this outcome may be attributed to the urban focus of our study and particularly the specific nutritional practices associated with the Newar ethnic group, who comprised majority of the sample. However, more research is essential to address the differences in prevalences.

A study in a rural population in India reported a similar prevalence of malnutrition as our study did [38].The study demonstrated a significant association between malnutrition and depression among the older people. These results align with existing literature, which has established a bidirectional relationship: depression serves as a risk factor for malnutrition, while poor nutritional status is conversely associated with an increased likelihood of depression [39].

In our study, malnutrition was positively associated with depression. This strong association suggests that a deficient nutritional state may act as a physiological stressor, ultimately triggering depressive symptoms. Consequently, malnutrition is identified not just as a comorbid condition, but as a significant independent risk factor for mental health decline in the older population [39].

Malnutrition and depression exist in a bidirectional, vicious cycle, where each condition exacerbates the other. Nutritional deficiencies directly cause depression through neurochemical changes, while depression causes malnutrition via loss of appetite, fatigue, or self-neglect [40,41,42]. This cycle is especially common in older populations and hospitalized patients. Compared with normally nourished individuals, malnourished individuals were more depressed [43,44].

Depressed individuals experience a loss of interest in work, decreased appetite, decreased self-care, and persistent sadness, which explains decreased feeding and malnutrition [45].

In addition, older adults ingest less food because of a greater degree of satiety due to decrease in gastric compliance, resulting in more rapid stomach filling and relaxation [46]. The enjoyment of food may be hampered in older adults and chronic inflammatory diseases common in older people are associated with anorexia [47].

Older individuals in the poor wealth quintile group may be more likely to be depressed than the very rich wealth quintile. Less wealthy participants may have increased risk of depression; an inverse relationship between wealth and depression, has been revealed in previous studies [48, 49]. However the association was seen statistically non significant in our study.

Additionally, female gender was significantly associated with depression as compared to their male counterparts. This sex disparity is consistent with the findings of other studies [24, 26, 50]. This may be due to more stressors and vulnerabilities that are prevalent in the lives of females than in the lives of males. Stressors cause negative mood, which leads to rumination. Females ruminate more than males do, which mediates the relationship between gender and depression [51]. The increased risk of depression in females compared with males highlights the need for targeted depression screenings and interventions in elderly females.

In this study, the older people who reported being financially dependent on others were significantly associated with depression. This can be explained by the fact that those with financial dependency on children may receive less care and support from family members, which is highly important in the prevention of depression [52].

A positive association was observed in those experiencing insomnia and depression. Insomnia is a risk factor for chronic pain and depression. Neuroinflammation plays a major role in the pathogenesis of both depression and chronic pain [53] and chronic pain causes insomnia. It is evidenced that insomnia is linked with depression and depression is associated with insomnia [54]. Additionally, current smoking was associated with depression in our study which is consistent with previous studies [55,56]. The plausible biological mechanisms to explain how tobacco smoke may cause depression have been described previously [57]. Smoking alters global brain activity influencing cognitive and emotional networks in an individual [56].

### Strengths and Limitations

This is a community-based study among the older adults in Nepal, that describes the status of depression, loneliness, and nutrition. A primary strength of this study lies in its use of validated assessment tools (GDS-15 and MNA), which ensures high data reliability and comparability. By investigating previously unmapped health dimensions in this community—specifically the factors associated with depression—this research fills a critical gap in Nepalese geriatric literature and establishes a baseline for future longitudinal studies and policy development. This study provides unique data on the intersection of mental health (depression and loneliness) and physical health (malnutrition) in a specific Nepalese context. Before this research, these dimensions had remained largely unexamined in this community, making this a vital baseline for regional health records.

Unlike studies that focus solely on clinical symptoms, this research adopted a comprehensive perspective by investigating the synergy between social isolation, psychological distress, and nutritional status. This multidimensional analysis offers a more complete picture of the challenges faced by older adults in Nepal. The findings offer actionable evidence for national and local program planners. By identifying specific “at-risk” population, the study provides a clear justification for targeted geriatric interventions and community-based mental health services.

Despite its strengths, this study has several limitations. The cross-sectional design limits our ability to draw causal inferences between depression and other variables. Another potential limitation of this study is recall bias, as the data relied on self-reported measures. Since the sample of the study is from urban areas of Nepal, findings from rural areas might vary. Important geriatric health screening tools, such as the Jong-Gierveld loneliness scale, and Lubben Social Network Scale, has not been validated previously in Nepal. It is important to conduct a validation study of the tool in the future in the Nepalese population. Finally, while the time gap between data collection and reporting is a limitation, the findings still provide a critical and previously undocumented valuable baseline information.

### Recommendations

To build upon these findings, future research should transition from cross-sectional to longitudinal designs to clarify the causal pathways between variables and mental health, loneliness and nutritional decline. Furthermore, integrating objective clinical markers alongside validated screening tools would also help mitigate recall bias and provide a more comprehensive overview of geriatric health in the community. To maintain geriatric wellbeing and prevent health decline, healthcare systems must become older age-friendly through specialized provider training and the systematic use of screening tools for depression, malnutrition, and social isolation. A holistic approach—integrating multidisciplinary rehabilitation, community mental health services, robust social networks along with fostering community-based social participation, and nutritional management—is essential for comprehensive old aged care. Implementing these grassroots measures is essential for preventing mental and physical decline. Given the high prevalence of depression, loneliness and malnutrition identified among the older people in Lalitpur, these findings emphasize the urgent need for focused mental health services, community-driven social engagement programs and nutritional support initiatives tailored to the older individuals of the community.

## Conclusions

This study highlights a high prevalence of depression, loneliness, and malnutrition among the older, with over 27% experiencing depression, more so in women than in men. Additionally, approximately half of the participants reported experiencing loneliness, and more than 58% suffered from poor nutritional status. Factors such as female sex, financial dependency, loneliness, insomnia, smoking status and malnutrition, as well as being at risk of malnutrition, were significantly associated with depression.

As population aging presents a growing societal challenge, these findings underscore the urgent need for public health measures. To ensure a high quality of life for older adults, strategies must be implemented to address mental health, social connectivity, and nutritional support systematically in the community.

A preprint of this manuscript has previously been published [58].

## Authors’ contributions

Conceptualization: Eebaraj Simkhada, Aerusha Simkhada Formal analysis: Eebaraj Simkhada

Investigation: Eebaraj Simkhada, Aerusha Simkhada Supervision: Eebaraj Simkhada.

Writing – original draft: Eebaraj Simkhada.

Writing – review & editing: Eebaraj Simkhada, Aerusha Simkhada

## Funding

This research did not receive any funding.

### Clinical trial number

Not applicable.

## Data availability

The dataset supporting the conclusions of this article is included within the article. The data are available upon request from the corresponding author.

## Declarations

### Ethics approval and consent to participate

This study was approved by the Institutional Review Committee, Institute of Medicine, Maharajgunj Medical Campus, Tribhuvan University, Nepal (Ref no. 75(6-11-E)071/072). This study followed the ethical standards laid down in the 1964 Declaration of Helsinki. All participants were informed of the study’s subject and method. Written consent was obtained from all participants, and thumb impressions were obtained from illiterate participants.

### Consent for publication

Not applicable.

## Data Availability

All data produced in the present study are available upon reasonable request to the corresponding author

## References

1. WHO (2015). WHO world report on Ageing 2015. 10.13140/RG.2.1.5058.8245.

2. National Population and Housing Census 2011 (National Report). Kathmandu, Nepal: Government of Nepal National Planning Commission Secretariat Central Bureau of Statistics, 2012.

3. Chen CC-H, Schilling LS, Lyder CH. A concept analysis of malnutrition in the elderly. Journal of Advanced Nursing. 2001;36: 131–142. 10.1046/j.13652648.2001.01950.x PMID: 11555057.

4. Status Report on Elderly People (60+) in Nepal on Health, Nutrition and Social Status Focusing on Research Needs. Kathmandu: Geriatric Center Nepal 2010.

5. Andrews G, Slade T, Sunderland M, Anderson T. Issues for DSM-V: Simplifying DSM-IV to Enhance Utility: The Case of Major Depressive Disorder. American Journal of Psychiatry. 2007;164: 1784–1785. 10.1176/appi.ajp.2007.07060928

6. Juruena MF, Eror F, Cleare AJ, Young AH. The Role of Early Life Stress in HPA Axis and Anxiety. Advances in Experimental Medicine and Biology. 2020;1191: 141–153. 10.1007/978-981-32-9705-0_9

7. Guigoz Y, Vellas B, Garry PJ. Assessing the nutritional status of the elderly: The Mini Nutritional Assessment as part of the geriatric evaluation. Nutr Rev. 1996; 54(1 Pt 2):S59±65. Epub 1996/01/01 PMID: 8919685.

8. Alamri SH, Bari AI, Ali AT. Depression and associated factors in hospitalized elderly: a cross-sectional study in a Saudi teaching hospital. Annals of Saudi Medicine. 2017;37: 122–129. 10.5144/0256-4947.2017.122

9. Chalise HN, Kai I, Saito T. Social Support and its Correlation with Loneliness: A Cross-Cultural Study of Nepalese Older Adults. The International Journal of Aging and Human Development. 2010;71: 115–138. 10.2190/ag.71.2.b

10. Nicholson NR. A review of social isolation: an important but underassessed condition in older adults. The journal of primary prevention. 2012;33: 137–52. 10.1007/s10935-012-0271-2

11. Cotterell N, Buffel T, Phillipson C. Preventing Social Isolation in Older People. Maturitas. 2018;113:80–4.

12. World Health Organization. Loneliness and isolation – the hidden threat to global health we can no longer ignore. 2025. https://www.who.int/news-room/commentaries/detail/loneliness-and-isolation-the-hidden-threat-to-global-health-we-can-no-longer-ignore. Accessed 10 December 2025.

13. Lee MR, Berthelot ER. Community Covariates of Malnutrition Based Mortality Among Older Adults. Annals of Epidemiology. 2010;20:371–9.

14. van Kan GA, Gambassi G, de Groot LCPGM, Andrieu S, Cederholm T, Andre E, et al. Nutrition and aging. The Carla workshop. The Journal of Nutrition Health and Aging. 2008;12:355–64.

15. FAO, IFAD and WFP. 2014. The State of Food Insecurity in the World 2014. Strengthening the enabling environment for food security and nutrition. Rome, FAO.

16. Chalise HN, Saito T, Kai I. Correlates of loneliness among older Newar adults in Nepal. Nihon Koshu Eisei Zasshi. 2007 Jul;54(7):427–33. PMID: 17763707.

17. Lyons G. Malnutrition: Elderly People in Nepal: Research briefing note. Kathmandu: Ageing Nepal University of Sheffield,. 2012.

18. Bakrania, S. (2015) Urban poverty in Nepal (GSDRC Helpdesk Research Report 1322) Birmingham, UK: GSDRC, University of Birmingham. Available from: https://gsdrc.org/wp-content/uploads/2016/01/HDQ1322.pdf

19. 19. Wikipedia Contributors. Lalitpur District. Wikipedia. 2020. https://en.wikipedia.org/wiki/Lalitpur_District,_Nepal. Accessed 10 December 2025.

20. Chalise HN, Saito T, Takahashi M, Kai I. Relationship specialization amongst sources and receivers of social support and its correlations with loneliness and subjective well-being: A cross sectional study of Nepalese older adults. Archives of Gerontology and Geriatrics. 2007;44:299–314.

21. Thapa P, Chakraborty P, Khattri J, Ramesh K, Sharma B. Psychiatric morbidity in elderly patients attending OPD of tertiary care centre in western region of Nepal. Industrial Psychiatry Journal. 2014;23:101.

22. Guigoz Y, Vellas B, Garry PJ. Assessing the Nutritional Status of the Elderly: The Mini Nutritional Assessment as Part of the Geriatric Evaluation. Nutrition Reviews. 2009;54:S59–65.

23. Kabir ZN, Ferdous T, Cederholm T, Khanam MA, Streatfied K, Wahlin Å. Mini Nutritional Assessment of rural elderly people in Bangladesh: the impact of demographic, socio-economic and health factors. Public Health Nutrition. 2006;9:968–74.

24. Ghimire S, Baral BK, Callahan K. Nutritional assessment of community-dwelling older adults in rural Nepal. PLOS ONE. 2017;12:e0172052.

25. Gautam R, Phd H. Geriatric Depression Scale for community-dwelling older adults in Nepal. Asian Journal of Gerontology & Geriatrics. 2011;6:93–102.

26. Boulos C, Salameh P, Barberger-Gateau P. The AMEL study, a cross sectional population-based survey on aging and malnutrition in 1200 elderly Lebanese living in rural settings: protocol and sample characteristics. BMC Public Health. 2013;13:573.

27. Yesavage JA, Brink TL, Rose TL, Lum O, Huang V, Adey M, et al. Development and Validation of a Geriatric Depression Screening scale: a Preliminary Report. Journal of Psychiatric Research. 1982;17:37–49.

28. Wilson RS, Krueger KR, Arnold SE, Schneider JA, Kelly JF, Barnes LL, et al. Loneliness and risk of Alzheimer disease. Archives of general psychiatry. 2007;64:234–40.

29. Lubben J, Blozik E, Gillmann G, Iliffe S, von Renteln Kruse W, Beck JC, et al. Performance of an Abbreviated Version of the Lubben Social Network Scale Among Three European Community-Dwelling Older Adult Populations. The Gerontologist. 2006;46:503–13.

30. Rutstein, Shea O. and Kiersten Johnson. The DHS Wealth Index. DHS Comparative Reports No. 6. Calverton, Maryland: ORC Macro. 2004.

31. Roodman, David Malin; MacKinnon, James G.; Nielsen, Morten Ørregaard; Webb, Matthew (2018): Fast and Wild: Bootstrap Inference in Stata Using boottest, Queen’s Economics Department Working Paper, No. 1406, Queen’s University, Department of Economics, Kingston (Ontario)

32. Penning MJ, Liu G, Chou PHB. Measuring Loneliness Among Middle-Aged and Older Adults: The UCLA and de Jong Gierveld Loneliness Scales. Social Indicators Research. 2013;118:1147–66.

33. World Health Organization. From loneliness to social connection-charting a path to healthier societies: report of the WHO Commission on Social Connection. 2025. https://iris.who.int. Accessed 10 December 2025.

34. Wojszel ZB. Determinants of nutritional status of older people in long-term care settings on the example of the nursing home in Białystok. Advances in medical sciences. 2006;51:168–73.

35. Locher JL, Ritchie CS, Roth DL, Baker PS, Bodner EV, Allman RM. Social isolation, support, and capital and nutritional risk in an older sample: ethnic and gender differences. Social Science & Medicine. 2005;60:747–61.

36. Holvast F, Burger H, de Waal MMW, van Marwijk HWJ, Comijs HC, Verhaak PFM. Loneliness is associated with poor prognosis in late-life depression: Longitudinal analysis of the Netherlands study of depression in older persons. Journal of Affective Disorders. 2015;185:1–7.

37. Vanessa Burholt, Thomas Scharf, Poor Health and Loneliness in Later Life: The Role of Depressive Symptoms, Social Resources, and Rural Environments, *The Journals of Gerontology: Series B*, Volume 69, Issue 2, March 2014, Pages 311–324, 10.1093/geronb/gbt121

38. Agarwalla R, Saikia A, Baruah R. Assessment of the nutritional status of the elderly and its correlates. Journal of Family and Community Medicine. 2015;22:39.

39. Rahe, C., Berger, K. (2016). Nutrition and Depression: Current Evidence on the Association of Dietary Patterns with Depression and Its Subtypes. In: Baune, B., Tully, P. (eds) Cardiovascular Diseases and Depression. Springer, Cham. 10.1007/978-3-319-32480-7_17

40. Sekhon S; Gupta V. Mood Disorder. 2023. https://pubmed.ncbi.nlm.nih.gov/32644337. Accessed 10 December 2025.

41. Sarris J, Logan AC, Akbaraly TN, Amminger GP, Balanzá-Martínez V, Freeman MP, Hibbeln J, Matsuoka Y, Mischoulon D, Mizoue T, Nanri A, Nishi D, Ramsey D, Rucklidge JJ, Sanchez- Villegas A, Scholey A, Su KP, Jacka FN; International Society for Nutritional Psychiatry Research. Nutritional medicine as mainstream in psychiatry. Lancet Psychiatry. 2015 Mar;2(3):271–4. doi: 10.1016/S2215-0366(14)00051-0. Epub 2015 Feb 25. PMID: 26359904.

42. German L, Feldblum I, Bilenko N, Castel H, Harman-Boehm I, Shahar DR. Depressive symptoms and risk for malnutrition among hospitalized elderly people. J Nutr Health Aging. 2008 May;12(5):313–8. doi: 10.1007/BF02982661. PMID: 18443713; PMCID: PMC12892648.

43. Gündüz E, Eskin F, Gündüz M, Bentli R, Zengin Y, Dursun R, İçer M, Durgun HM, Gürbüz H, Ekinci M, Yeşil Y, Güloğlu C. Malnutrition in Community-Dwelling Elderly in Turkey: A Multicenter, Cross-Sectional Study. Med Sci Monit. 2015 Sep 15;21:2750–6. doi: 10.12659/MSM.893894. PMID: 26371941.

44. Wei J, Fan L, Zhang Y, et al. Association Between Malnutrition and Depression Among Community-Dwelling Older Chinese Adults. Asia Pacific Journal of Public Health. 2018;30(2):107–117. doi:10.1177/1010539518760632

45. Ülger Z, Halil M, Kalan I, Yavuz BB, Cankurtaran M, Güngör E, et al. Comprehensive assessment of malnutrition risk and related factors in a large group of community-dwelling older adults. Clinical Nutrition. 2010;29:507–11.

46. MacIntosh, C, Morley JE, Chapman IM. The anorexia of aging. Nutrition. 2000.

47. Francesco VD, Fantin F, Omizzolo F, Residori L, Bissoli L, Bosello O, et al. The Anorexia of Aging. Digestive Diseases. 2007;25:129–37.

48. Osafo Hounkpatin, H., Wood, A.M., Brown, G.D.A., et al. Why does Income Relate to Depressive Symptoms? Testing the Income Rank Hypothesis Longitudinally. Soc Indic Res 124, 637–655 (2015). 10.1007/s11205-014-0795-3

49. Brinda EM, Rajkumar AP, Attermann J, Gerdtham UG, Enemark U, Jacob KS. Health, Social, and Economic Variables Associated with Depression Among Older People in Low and Middle Income Countries: World Health Organization Study on Global AGEing and Adult Health. Am J Geriatr Psychiatry. 2016 Dec;24(12):1196–1208. doi: 10.1016/j.jagp.2016.07.016. Epub 2016 Jul 25. PMID: 27743841.

50. AlbertPaul R.. 2015. Why is depression more prevalent in women?. Journal of Psychiatry and Neuroscience. 40(4): 219–221. 10.1503/jpn.150205

51. Girgus J, Yang K, Ferri C. The Gender Difference in Depression: Are Elderly Women at Greater Risk for Depression Than Elderly Men? Geriatrics. 2017;2:35.

52. Buvneshkumar M, John KR, Logaraj M. A study on prevalence of depression and associated risk factors among elderly in a rural block of Tamil Nadu. Indian Journal of Public Health. 2018;62:89–94.

53. Zis P, Daskalaki A, Bountouni I, Sykioti P, Varrassi G, Paladini A. Depression and chronic pain in the elderly: links and management challenges. Clinical Interventions in Aging. 2017;Volume 12:709–20

54. Fang H, Tu S, Sheng J, Shao A. Depression in sleep disturbance: A review on a bidirectional relationship, mechanisms and treatment. J Cell Mol Med. 2019 Apr;23(4):2324–2332. doi: 10.1111/jcmm.14170. Epub 2019 Feb 7. PMID: 30734486; PMCID: PMC6433686.

55. Almeida OP, Pfaff JJ. Depression and smoking amongst older general practice patients. J Affect Disord. 2005 Jun;86(2-3):317–21. doi: 10.1016/j.jad.2005.02.014. PMID: 15935254.

56. Velioglu HA, Yıldız S, Ozdemir-Oktem E, Cankaya S, Lundmark AK, Ozsimsek A, Hanoglu L, Yulug B. Smoking affects global and regional brain entropy in depression patients regardless of depression: Preliminary findings. J Psychiatr Res. 2024 Sep;177:147–152. doi: 10.1016/j.jpsychires.2024.07.002. Epub 2024 Jul 3. PMID: 39018709.

57. Taylor AE, Fluharty ME, Bjørngaard JH, et al. Investigating the possible causal association of smoking with depression and anxiety using Mendelian randomisation meta-analysis: the CARTA consortium. BMJ Open. 2014;4(10):e006141. Published 2014 Oct 7. doi:10.1136/bmjopen-2014-006141

58. Eebaraj Simkhada, Aerusha Simkhada. Depression, loneliness, and poor nutrition among elderly people in Nepal: A cross-sectional study, 2026, PREPRINT (Version 1) available at Research Square [10.21203/rs.3.rs-8329238/v1]

